# Influence of alcohol-serving venue characteristics on yield of HIV status-neutral screening in rural East Africa

**DOI:** 10.1101/2025.08.01.25332709

**Authors:** Janice Litunya, Brian Beesiga, Joy Z. Nakato, Jaquiline Mwango, Kara Marson, Erick Wafula Mugoma, Jennifer Temple, Pius Agaba, Jane Kabami, Elijah Kakande, Judith A. Hahn, Carol S. Camlin, Starley B. Shade, Maya L. Petersen, Diane V. Havlir, Moses R. Kamya, Laura B. Balzer, James Ayieko, Gabriel Chamie

**Author notes:** Corresponding author:, (JL).

## Abstract

Alcohol-serving venues are high-risk sites for HIV transmission in East Africa. Understanding how venue characteristics influence HIV screening outcomes may help to target venue-based outreach. In eight rural communities in Kenya and Uganda, we mapped all alcohol-serving venues (N=530) and invited owners to participate in a cluster-randomized trial to promote biomedical HIV prevention uptake: 527 (99%) owners agreed to participate. We distributed cards recruiting adults ( ≥ 18 years) for free HIV testing with rapid initiation of HIV biomedical prevention or treatment. We characterized the yield of venue-based recruitment and evaluated venue-level correlates of being newly diagnosed with HIV, being previously diagnosed but out-of-care, and having self-reported HIV risk. Of 480 participating venues (Kenya=89, Uganda=391; 41 closed pre-recruitment and 6 had no one present), 61 (13%) had rooms for sex work; 91 (19%) offered condoms, and the median patrons/venue was 10/weekend-day. Staff distributed 9,375 cards and 7,744 (83%) adults participated in HIV screening. Of those screened, the median age was 34 years (IQR:26-43), 62% were men, and 1,620 (21%) had HIV. Among persons without known HIV, 141/6,265 (2.3%) were newly-diagnosed with HIV. Among persons with known HIV, 78/1,479 (5.3%) were out-of-care. Among persons without HIV, 2,285/6,124 (37%) reported HIV risk. The odds of having newly-diagnosed HIV increased significantly with each additional patron/weekend-day at a given venue (adjusted odds ratio [aOR]=1.03, 95%CI:1.00-1.05, p=0.025). The odds of being previously diagnosed but out-of-care were significantly lower among attendees at venues with condoms on site (aOR=0.39, 95%CI:0.16-0.99, p=0.047). The odds of reporting HIV risk were significantly higher among attendees at venues with condoms (aOR=1.25, 95%CI:1.04-1.49, p=0.015), more patrons/weekday (aOR=1.01, 95%CI:1.00-1.02, p=0.022), and more barmaids (aOR=1.07, 95%CI:1.01-1.13, p=0.013). Alcohol-serving venue characteristics were predictive of the yield of persons with untreated HIV or high HIV risk, and could aid programs in targeting venues for HIV prevention and treatment.

## Introduction

Alcohol is the most widely used substance in Sub-Saharan Africa,[1] and associations between alcohol use and HIV risk behavior, incidence, and poor HIV care cascade outcomes are well documented. Alcohol use has been shown to increase HIV risk behavior by altering judgment and decision-making, thereby reducing one’s perception of risk.[2] Resultant high-risk sexual behaviors include having multiple sexual partners, inconsistent or no condom use and transactional sex.[3] The risk of HIV acquisition rises with alcohol consumption, with unhealthy alcohol use increasing the likelihood of HIV acquisition by two to five times compared to no alcohol use.[4] Despite this heightened risk, persons with unhealthy alcohol use globally are less likely to engage and maintain engagement with HIV prevention services compared to persons with no alcohol use.[5,6] In addition, several studies have shown that alcohol use negatively impacts HIV care cascade outcomes, with persons with HIV and unhealthy alcohol use having lower HIV status awareness,[7] antiretroviral treatment (ART) uptake and adherence,[8] and ultimately, lower HIV viral suppression.[9] As such, alcohol use is a driver of HIV transmission through the combined effects of reduced prevention use among persons at risk for HIV, worse care cascade outcomes among persons with HIV, and increased HIV transmission risk behaviors among both persons with and without HIV.

Alcohol-serving venues are sites of increased HIV transmission risk and offer opportunities to engage persons with and without HIV in community-based HIV screening for prevention and treatment.[10,11] In sub-Saharan Africa, these venues include neighborhood bars, night clubs, karaoke bars, taverns, wine shops, lounges, and informal venues such as home-based alcohol-serving venues. They serve diverse functions, providing spaces for recreation, socially accepted alcohol use, social networking, and opportunities to meet sexual partners.[10,12] Therefore, the settings in which people consume alcohol may influence alcohol-related sexual risks. In addition, studies have shown that both patrons and workers at alcohol-serving venues are at increased risk of HIV,[10] and that attendance at these venues is associated with increased risk of HIV, independent of alcohol consumption.[13] Additionally, a recent national cross-sectional survey of social venues in Uganda found increased self-report of HIV vulnerability among young venue workers at venues serving alcohol and with on-site sex work, as compared to venues without these characteristics.[14]

Although alcohol-serving venues are known HIV transmission hubs, there remains limited understanding of whether and how their characteristics are associated with HIV screening outcomes in East Africa. To address this gap, we recruited adults (≥18 years) from alcohol-serving venues in rural Kenya and Uganda and evaluated the associations between venue-level characteristics and linking to a health facility for HIV screening, having untreated HIV, or being at increased risk of HIV infection.

## Methods

### Study design and setting

We evaluated associations between characteristics of alcohol-serving venues and HIV screening outcomes using cross-sectional data obtained from a cluster randomized trial of mobilization strategies to promote biomedical HIV prevention uptake among adults recruited from alcohol-serving venues in rural Kenya and Uganda (OPAL-East Africa trial: NCT05862857). The study was carried out in eight communities of ∼10,000 residents per community in western Kenya and southwestern Uganda. Each community is within the catchment area of a government-associated health center providing HIV care, including ART and biomedical HIV prevention services, including pre-exposure prophylaxis (PrEP) and post-exposure prophylaxis (PEP).

### Procedures and study population

We worked with local community leaders to map all alcohol-serving venues in the study communities. These venues were defined as any community locations where alcohol was sold and consumed, including bars, nightclubs, hotels and informal drinking dens. We sought permission from venue owners to recruit from their venues as part of the study. At venues in which owners agreed to participate, we measured venue characteristics using a standardized venue survey. We classified venues as “formal” if licensed to sell alcohol (including bars, nightclubs or restaurants/guesthouses) and “informal” if unlicensed (such as households and outdoor areas like beaches). Study staff interviewed venue owners/managers during venue visits and recorded the number of rooms in the venue, the estimated number of patrons per weekend-day and weekday, number of workers, number of barmaids (the subset of venue workers who serve alcohol), availability of rooms for commercial sex work, type of alcohol sold (i.e., commercially-vs locally-produced alcohol or both), presence of indoor and/or outdoor alcohol-serving spaces, and availability of condoms on site, as reported by the venue owner or manager. As part of the OPAL-East Africa trial, we created clusters of geographically nearby venues and randomized them to receive recruitment cards for multi-disease screening (including HIV, hypertension, diabetes, malaria, TB disease, pregnancy and syndromic sexually-transmitted infection screening [intervention]) or HIV-only screening (control) at the nearby health facility.

From 13 July 2023 to 10 December 2024, research staff conducted two visits per venue, targeting the busiest times of venue activity as identified by the venue owners, to recruit both patrons and venue workers. During each venue visit, study staff spoke with adult patrons and workers, encouraged them to come to the local clinic for further health screening, and distributed recruitment cards. Staff recorded the sex, the estimated age, and country of the person who received the recruitment card.

Individuals who went to the facility with a recruitment card and were ≥18 years of age were eligible to participate in health facility-based HIV screening. Individuals who were <18 years old or unable to provide consent (e.g. if heavily intoxicated from alcohol use at the time of arriving at the facility) were not eligible for study participation. Eligible adults who provided written informed consent were enrolled in the study, completed a socio-demographic survey, and underwent HIV screening. Participants who did not report a known HIV diagnosis were asked about self-reported current, or anticipated future (next 3-month) HIV risk, and underwent HIV testing by rapid antibody testing, according to national (Kenya or Uganda) Ministry of Health guidelines.[15,16] Participants who reported a prior HIV diagnosis were asked about use of ART and engagement in HIV care, but not retested. Participants with newly-diagnosed HIV and participants who reported known HIV but were out-of-care or not on ART were referred for same-day ART initiation (or re-initiation) at the local health facility. Participants who tested HIV negative were referred for same-day biomedical HIV prevention services, including PrEP or PEP, at the local health facility. Participants who screened positive for other conditions in the multi-disease screening group were referred for same-day, further evaluation and/or treatment at the local health facility.

Clinic-based screening and ART or PrEP/PEP services were offered free of charge. Staff obtained digital fingerprint biometric measures at enrollment, and cross-checked fingerprint biometrics over the course of the study to ensure participants were only enrolled once. All participants who were enrolled received a travel reimbursement of ∼US$5 at the end of their visit to the health facility. Individuals who presented more than one time for screening were referred for services at the health facility but were not re-enrolled and were not provided additional travel reimbursements.

### Outcomes

We determined whether or not a recruitment card resulted in a person coming to the health facility for screening. We also classified outcomes among enrolled persons who participated in screening as: (i) newly-diagnosed HIV (i.e., HIV rapid antibody positive), (ii) with self-reported known HIV but out-of-care, or (iii) HIV-uninfected with self-reported increased risk of HIV infection (current or anticipated risk in the next three months). We considered persons with newly-diagnosed HIV or with known HIV but out-of-care as having untreated HIV. The main study outcomes were yield of adults who participated in HIV screening following venue-based recruitment, and among persons who participated in HIV screening, yield of being newly diagnosed with HIV, being previously diagnosed but out-of-care, or being HIV-uninfected and having self-reported HIV risk.

### Statistical Analysis

We described alcohol-serving venue characteristics and the proportion of recruitment cards distributed that resulted in persons given cards at those venues subsequently attending health facility-based screening. Then we used Generalized Estimating Equations (GEE) to investigate associations between venue characteristics and the yield of screening.[17] Venue characteristics included: formal/informal, type of alcohol sold (i.e., commercially-vs locally-produced alcohol or both), number of rooms in the venue, presence of indoor and/or outdoor alcohol-serving spaces, estimated number of patrons per weekend-day and weekday, number of workers, number of barmaids, availability of rooms for commercial sex work, and availability of condoms on site. We first evaluated venue-level predictors of participating in facility-based screening. Then we evaluated venue-level predictors of the following health outcomes among those screened: newly-diagnosed with HIV, previously diagnosed with HIV but out-of-care, or HIV-uninfected with self-reported HIV risk. We accounted for the correlation by drinking venue with an independent working correlation matrix. The primary analyses adjusted for age, sex and country and resulted in adjusted odds ratios (aOR). Secondary analyses were unadjusted odds ratios of the same outcomes. All analyses were pooled over trial arm and were conducted using R version 4.4.2.

### Ethical Approval

The Kenya Scientific and Ethics Review Unit, the Kenya National Commission for Science, Technology and Innovation, the Makerere University School of Medicine Research and Ethics Committee in Uganda, the Uganda National Council for Science and Technology, and the University of California San Francisco Committee on Human Research in the United States, reviewed and approved the study protocol. All participants provided written informed consent prior to study participation.

## Results

In eight communities, study staff mapped a total of 530 alcohol-serving venues, of which 527 (99%) venues (i.e., owners) agreed to participate in the trial: 93 venues in Kenya and 434 in Uganda. From July 2023 to January 2025, staff visited these venues to distribute recruitment cards. Prior to receiving a visit, 41 (8%) closed (3=Kenya and 38=Uganda). Six (1%) venues were visited twice but no one was present during visits (1=Kenya and 5=Uganda). Overall, 480 venues were open and had patrons or workers present for recruitment (89 venues in Kenya and 391 venues in Uganda).

Characteristics of participating venues are shown in **Table 1**. Though a greater number of venues were enrolled in Uganda, the median number of patrons (e.g. 15 vs 8 patrons/weekend-day) and workers (2 vs. 1 workers) per venue were larger in Kenya than Uganda, respectively. Overall, 426/480 (89%) of the venues were informal; 153 (32%) venues sold locally-produced alcohol only; 72 (15%) sold commercially-produced alcohol only, and 255 (53%) sold both. The median number of patrons was 8 per weekday and 10 per weekend-day. Sixty-one (13%) venues had rooms for sex work (4% in Kenya vs 15% in Uganda), and 91 (19%) venues offered condoms on site (3% in Kenya vs 23% in Uganda). Photographs of several venue characteristics are shown in **Fig 1**.

**Fig 1.**
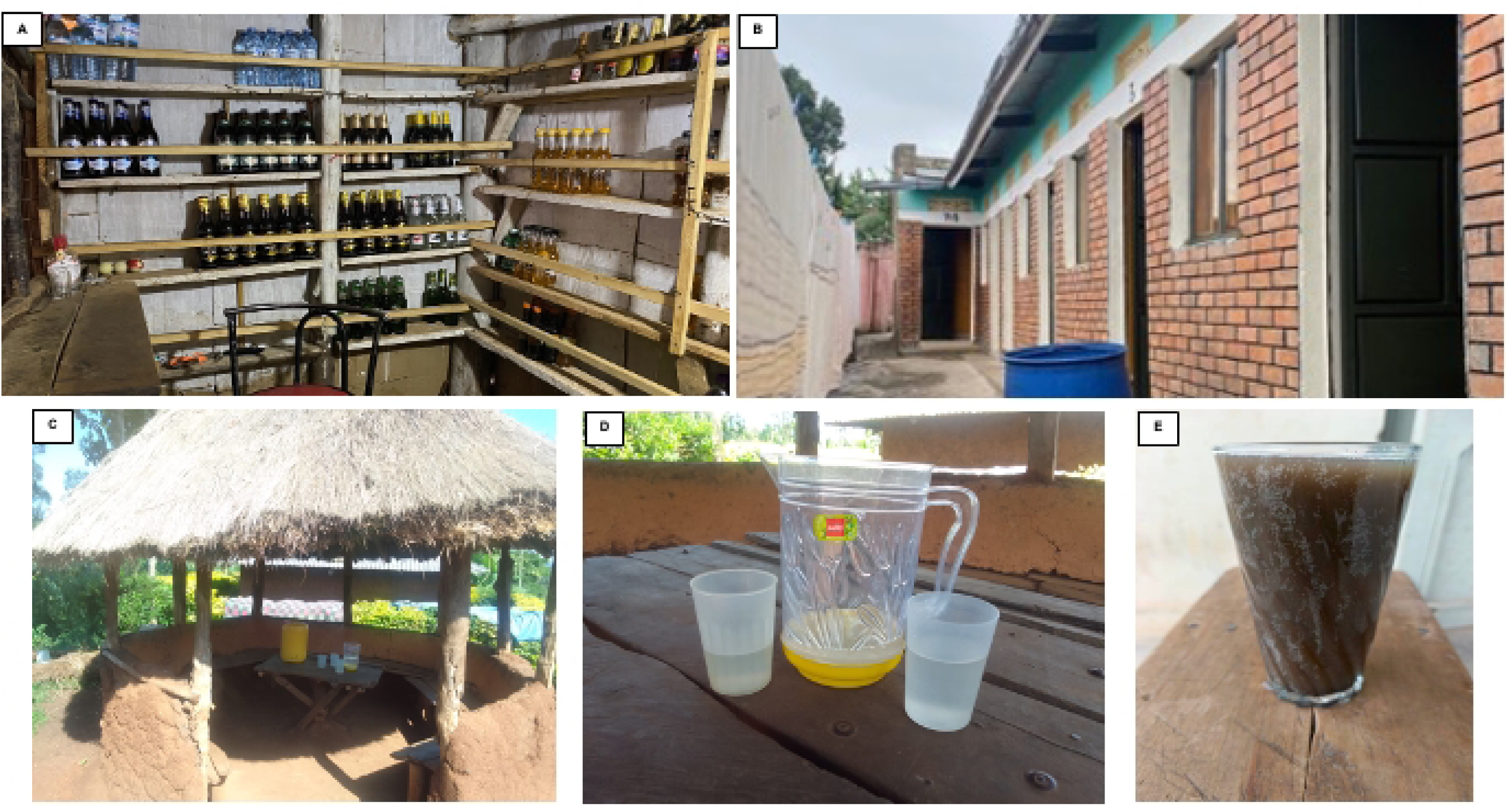
Photos of alcohol-serving venue characteristics in Kenya and Uganda. (A) Commercial alcohol at a venue in Uganda; (B) Rooms for sex work at a venue in Uganda; (C) Outdoor, informal venue in Kenya; (D) Locally-produced alcohol at a venue in Kenya; (E) Locally-produced alcohol at a venue in Uganda.

**Table 1:** Characteristics of alcohol-serving venues from which patrons and workers were recruited for clinic-based HIV screening in eight communities in rural Kenya and Uganda.

| Alcohol-serving venue characteristics | Kenyan Venues<br>(N=89) | Ugandan Venues<br>(N=391) | Total<br>(N=480) |
| --- | --- | --- | --- |
| *Venue type\** |  |  |  |
| Formal | 17 (19%) | 37 (9%) | 54 (11%) |
| Informal | 72 (81%) | 354 (91%) | 426 (89%) |
| *Type of alcohol sold* |  |  |  |
| Locally made alcohol only | 67 (75%) | 86 (22%) | 153 (32%) |
| Commercially made alcohol only | 21 (24%) | 51 (13%) | 72 (15%) |
| Both locally made & commercial alcohol | 1 (1%) | 254 (65%) | 255 (53%) |
| Number of rooms per venue, median (range) | 1 (1-6) | 1 (1-10) | 1 (1-10) |
| *Bar area\*\** |  |  |  |
| Indoor | 18 (20%) | 161 (42%) | 179 (38%) |
| Predominantly indoor | 5 (6%) | 129 (34%) | 134 (28%) |
| Evenly mixed indoor and outdoor | 5 (6%) | 38 (10%) | 43 (9%) |
| Predominantly outdoor | 31 (35%) | 28 (7%) | 59 (13%) |
| Outdoor | 5 (6%) | 38 (19%) | 43 (9%) |
| Patrons per weekday, median (IQR) | 10 (8,18) | 8 (5,12) | 8 (5,15) |
| Patrons per weekend, median (IQR) | 15 (10,20) | 8 (5,12) | 10 (5,15) |
| Number of venue workers, median (IQR) | 2 (1,2) | 1 (1,2) | 1 (1,2) |
| Number of barmaids, median (IQR) | 1 (0,2) | 1 (1,1) | 1 (1,1) |
| Venues with rooms for sex work | 4 (4%) | 57 (15%) | 61 (13%) |
| Venues with condoms on site** | 3 (3%) | 88 (23%) | 91 (19%) |

\*Formal alcohol-serving venues include legal, designated stand-alone bars, and night clubs, restaurants and hotels/guesthouses with associated bars. Informal venues include households, "drinking dens" and outdoor sites (such as beaches or fields).
\*\*Data on bar area and condom availability missing for nine venues: 1 from Kenya and 8 from Uganda.
IQR: Interquartile range

Demographic characteristics of venue attendees (patrons and workers) recruited are shown in **Table 2**. Study staff distributed a total of 9,375 recruitment cards: 4,596 in Kenya and 4,779 in Uganda. Overall, staff distributed 6,031 cards (64%) to men and 3,344 (36%) to women. Of 8,670 cards with recorded patron/worker status, staff distributed cards to 7,803 (90%) patrons and 867 (10%) workers.

**Table 2:** Age, sex and patron/worker status of adults (≥18 years of age) recruited from alcohol-serving venues in eight study communities in rural Kenya and Uganda.

|  | Kenya<br>N=4596 adults | Uganda<br>N=4779 adults | Total<br>N=9375 adults |
| --- | --- | --- | --- |
| Male | 2907 (63%) | 3124 (65%) | 6031 (64%) |
| Age, median (IQR) | 32 (26-41) | 32 (26,42) | 32 (26,42) |
| Patron* | 4001/4164 (96%) | 3802/4506 (84%) | 7803/8670 (90%) |
| Worker* | 163/4164 (4%) | 704/4506 (16%) | 867/8670 (10%) |

\*Patron/worker status missing for 705 adults (432 adults in Kenya and 273 adults in Uganda).

Overall, 7,744 (83%) individuals who received recruitment cards came to the health facility for clinic-based HIV screening and were enrolled. Several venue characteristics were associated with significantly lower recruitment yield **(Table 3)**. Specifically, the odds of participating in facility-based screening were significantly lower for persons recruited from venues that were, formal (aOR: 0.74, 95%CI: 0.61-0.90, p=0.003), venues that served commercial alcohol only (aOR: 0.56, 95%CI: 0.46-0.68, p<0.001; reference=locally-produced alcohol only), venues with more rooms (aOR: 0.82, 95% confidence interval [CI]: 0.78-0.87, p<0.001, per additional room), venues with more patrons (aOR: 0.98, 95%CI: 0.98-0.99, p<0.001, per additional patron), venues with more workers (aOR: 0.90, 95%CI: 0.87-0.94,p<0.001, per additional worker), venues with more barmaids (aOR=0.90, 95%CI: 0.86-0.94, p<0.001, per additional barmaid), venues with rooms for sex work (aOR: 0.52, 95%CI: 0.44-0.62, p<0.001) and with condoms on-site (aOR: 0.76, 95%CI: 0.65-0.89, p=0.001). As compared to venues that were entirely indoor, attending the health clinic screening was more likely when recruitment was from venues that were outdoors (aOR: 1.34, 95% CI: 1.04-1.73, p=0.023) or mixed but predominantly outdoors (aOR: 1.35, 95% CI: 1.05-1.72, p=0.018), but attendance was lower when recruiting from venues that had an even mix of indoor and outdoor spaces (aOR: 0.78, 95%CI: 0.61-0.99, p=0.042). Unadjusted analyses yielded similar results (Supplementary materials: **S1 Table)**.

**Table 3:** Venue characteristics associated with likelihood of linking to facility-based screening among participants who received recruitment cards, with odds ratios adjusted for age, sex and country.

| Venue characteristics | aOR | 95% CI | p-value |
| --- | --- | --- | --- |
| Formal bar (vs. informal venue) | 0.74 | 0.61 - 0.90 | **0.003** |
| Venue serving commercial brew only | 0.56 | 0.46 - 0.68 | **<0.001** |
| Venue serving both local and commercial brew | 0.86 | 0.72 - 1.04 | 0.116 |
| Number of rooms (per 1 room increase) | 0.82 | 0.78 - 0.87 | **<0.001** |
| Outdoor venues | 1.34 | 1.04 - 1.73 | **0.023** |
| Mixed but predominantly indoor | 1.10 | 0.93 - 1.31 | 0.257 |
| Mixed but predominantly outdoor | 1.35 | 1.05 - 1.72 | **0.018** |
| Evenly mixed indoor and outdoor | 0.78 | 0.61 - 0.99 | **0.042** |
| Patrons per weekday (per 1 patron increase) | 0.98 | 0.98 - 0.99 | **<0.001** |
| Patrons per weekend (per 1 patron increase) | 0.99 | 0.98 - 0.99 | **<0.001** |
| Venue workers (per 1 worker increase) | 0.90 | 0.87 - 0.94 | **<0.001** |
| Venue barmaid (per 1 barmaid increase) | 0.90 | 0.86 - 0.94 | **<0.001** |
| Rooms for sex work | 0.52 | 0.44 - 0.62 | **<0.001** |
| Condoms available on site | 0.76 | 0.65 - 0.89 | **0.001** |

Characteristics of recruited adults who came to the health facility and participated in screening are shown in **Table 4**. Of 7,774 participants, their overall median age was 34 years (interquartile range [IQR]: 26-43 years), 62% were men, median daily was US$2 (IQR:$1-3), 54% were married and living with their spouse, and 91% were venue patrons. The median AUDIT-C score was 4 (IQR: 2-7) in Uganda and 3 (IQR: 0-5) in Kenya. Of adults who linked for screening, HIV positivity was 21% (1,620/7,744): 25% in Kenya and 16% in Uganda. Among persons without known HIV prior to screening, 141/6265 (2.3%) were newly diagnosed with HIV. The yield of persons with newly-diagnosed HIV was higher in Uganda at 3.3% (107/3,203) than Kenya at 1.1% (34/3,062). Among persons with previously diagnosed HIV, 78/1,479 (5.3%) individuals reported being out of HIV care. Among persons who tested HIV negative, 1253/3028 (41%) and 1032/3096 (33%) reported increased HIV risk in Kenya and Uganda respectively **(Table 4).**

**Table 4:**
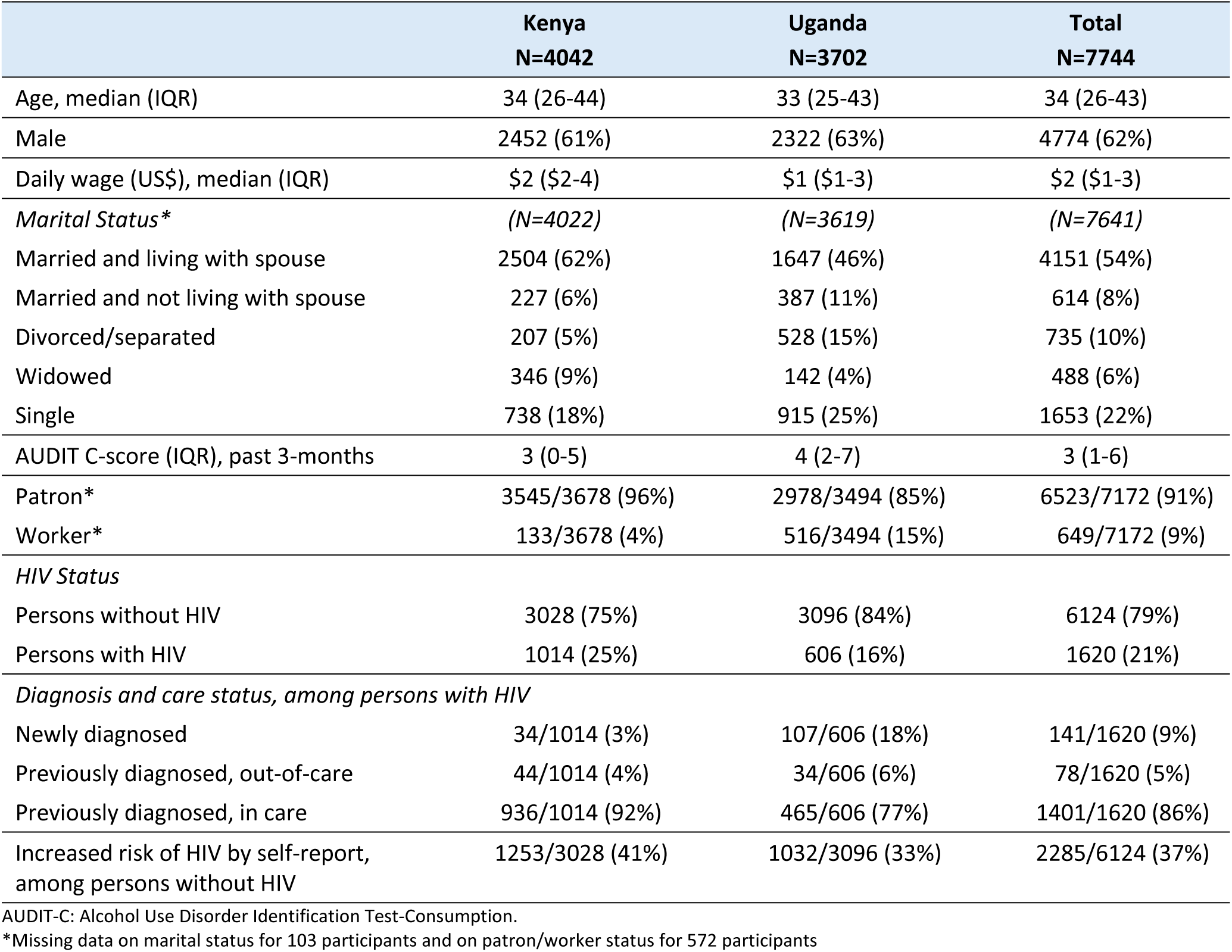
Characteristics of adults who linked to health facilities and participated in HIV screening following recruitment from alcohol-serving venues in eight communities in rural Kenya and Uganda.

|  | Kenya<br>N=4042 | Uganda<br>N=3702 | Total<br>N=7744 |
| --- | --- | --- | --- |
| Age, median (IQR) | 34 (26-44) | 33 (25-43) | 34 (26-43) |
| Male | 2452 (61%) | 2322 (63%) | 4774 (62%) |
| Daily wage (US\$), median (IQR) | \$2 (\$2-4) | \$1 (\$1-3) | \$2 (\$1-3) |
| *Marital Status\** | (N=4022) | (N=3619) | (N=7641) |
| Married and living with spouse | 2504 (62%) | 1647 (46%) | 4151 (54%) |
| Married and not living with spouse | 227 (6%) | 387 (11%) | 614 (8%) |
| Divorced/separated | 207 (5%) | 528 (15%) | 735 (10%) |
| Widowed | 346 (9%) | 142 (4%) | 488 (6%) |
| Single | 738 (18%) | 915 (25%) | 1653 (22%) |
| AUDIT C-score (IQR), past 3-months | 3 (0-5) | 4 (2-7) | 3 (1-6) |
| Patron* | 3545/3678 (96%) | 2978/3494 (85%) | 6523/7172 (91%) |
| Worker* | 133/3678 (4%) | 516/3494 (15%) | 649/7172 (9%) |
| *HIV Status* |  |  |  |
| Persons without HIV | 3028 (75%) | 3096 (84%) | 6124 (79%) |
| Persons with HIV | 1014 (25%) | 606 (16%) | 1620 (21%) |
| *Diagnosis and care status, among persons with HIV* |  |  |  |
| Newly diagnosed | 34/1014 (3%) | 107/606 (18%) | 141/1620 (9%) |
| Previously diagnosed, out-of-care | 44/1014 (4%) | 34/606 (6%) | 78/1620 (5%) |
| Previously diagnosed, in care | 936/1014 (92%) | 465/606 (77%) | 1401/1620 (86%) |
| Increased risk of HIV by self-report, among persons without HIV | 1253/3028 (41%) | 1032/3096 (33%) | 2285/6124 (37%) |

AUDIT-C: Alcohol Use Disorder Identification Test-Consumption.
\*Missing data on marital status for 103 participants and on patron/worker status for 572 participants

In multivariate analyses, the odds of being newly-diagnosed with HIV increased significantly with each additional patron per weekend-day at a given venue (aOR: 1.03, 95%CI: 1.00-1.05, p=0.025). The odds of being previously diagnosed with HIV, but out-of-care were significantly lower among attendees at venues with condoms available on-site (aOR: 0.39, 95% CI: 0.16-0.99, p=0.047). The odds of reporting increased HIV risk were significantly higher among attendees at venues with a greater number of patrons per weekday, a greater number of barmaids, and with condoms available on site (**Table 5)**. In unadjusted analyses, the odds of being newly-diagnosed with HIV were increased for several additional venue characteristics, including selling commercial alcohol, having a greater number of rooms, having rooms for sex work and having condoms available on-site; the odds of being newly-diagnosed with HIV were decreased with outdoor venues (Supplementary materials, **S2 Table**).

**Table 5:** Associations between alcohol-serving venue characteristics and HIV screening outcomes among adults recruited from alcohol-serving venues, including: a) newly diagnosed HIV, b) known HIV and out-of-care, and c) increased risk of HIV, if HIV uninfected; with adjusted odds ratios (aOR) controlling for age, sex and country.

| Venue characteristics | a) Newly diagnosed HIV<br>aOR (95% CI)<br>p-value | b) Known HIV+, out-of-care<br>aOR (95% CI)<br>p-value | c) Increased HIV risk, if HIV-<br>aOR (95% CI)<br>p-value |
| --- | --- | --- | --- |
| Formal bar (vs. informal venue) | 1.33 (0.79 - 2.22)<br>p=0.280 | 0.54 (0.23 - 1.24)<br>p=0.147 | 0.86 (0.66 - 1.11)<br>p=0.240 |
| Venue serves commercial alcohol only (vs. locally made alcohol only) | 1.56 (0.93 - 2.63)<br>p=0.092 | 0.85 (0.44 - 1.67)<br>p=0.647 | 0.95 (0.72 - 1.24)<br>p=0.693 |
| Number of rooms (aOR per 1 room increase) | 1.17 (0.97 - 1.42)<br>p=0.102 | 1.04 (0.86-1.25)<br>p=0.686 | 1.02 (0.93-1.12)<br>p=0.647 |
| Patrons per weekday (aOR per 1 patron increase) | 1.01 (1.00 - 1.03)<br>p=0.120 | 1.00(0.98 - 1.02)<br>p=0.969 | 1.01 (1.00 - 1.02)<br>**p=0.022** |
| Patrons per weekend (aOR per 1 patron increase) | 1.03 (1.00 - 1.05)<br>**p=0.025** | 1.01 (0.98 - 1.04)<br>p=0.447 | 1.00 (0.98- 1.01)<br>p=0.416 |
| Venue workers (aOR per 1 worker increase) | 1.08 (0.94 - 1.23)<br>p=0.296 | 0.87 (0.70 - 1.07)<br>p=0.186 | 1.05 (0.99 - 1.11)<br>p=0.137 |
| Venue barmaid (aOR per 1 barmaid increase) | 1.10 (0.95 - 1.27)<br>p=0.196 | 0.79 (0.61 - 1.02)<br>p=0.067 | 1.07 (1.01 - 1.13)<br>**p=0.013** |
| Rooms for sex work | 1.24 (0.74 - 2.07)<br>p=0.416 | 0.99(0.45-2.18)<br>p=0.985 | 1.19 (0.97-1.46)<br>p=0.104 |
| Condoms available on site | 1.06 (0.65 – 1.71)<br>p=0.824 | 0.39 (0.16 – 0.99)<br>**p=0.047** | 1.25 (1.04 - 1.49)<br>**p=0.015** |

## Discussion

In this study we found that specific characteristics of alcohol-serving venues were associated with yield of adults who participated in HIV screening, and with the screening outcomes of newly diagnosed HIV, known HIV and out-of-care, and increased HIV risk, following venue-based outreach and referral to clinic-based HIV screening. Overall, formal venues, larger venues (in terms of rooms, patrons and workers) and venues with rooms for sex work and condoms available on-site were associated with significantly lower yield of adults who participated in HIV screening. Having a larger number of patrons per weekend-day was significantly associated with higher odds of identifying venue attendees with newly-diagnosed HIV, and having a larger number of patrons per weekday, a larger number of barmaids and condoms available on site were significantly associated with higher odds of identifying persons at increased risk of HIV, in multivariate analyses. In contrast, having condoms available on-site was associated with significantly lower odds of identifying persons previously diagnosed with HIV but out-of-care. Furthermore, in unadjusted analyses, our findings suggest that venues that serve commercial alcohol, and have more rooms, condoms available on site and rooms for sex work on site were associated with higher odds of identifying persons with undiagnosed HIV. These findings could aid programs in targeting venues for HIV prevention and treatment.

Alcohol-serving venues are known hubs of HIV transmission in rural sub-Saharan Africa due to a combination of unhealthy alcohol use and increased sexual risk behavior (including transactional sex) among persons at increased risk of both HIV acquisition and onward transmission (i.e., from persons with untreated HIV).[18] By providing an improved understanding of how alcohol-serving venue characteristics in rural sub-Saharan Africa relate to yield of adults at high risk of HIV transmission, these findings may aid in targeting a subset of alcohol-serving venues when conducting community-based outreach for HIV testing, prevention and treatment services.

Our findings add to the literature on the relationship between alcohol-serving venue characteristics and factors that increase HIV transmission risk. Prior research has shown that certain characteristics of alcohol-serving venues in sub-Saharan Africa, such as on-site sex work in Uganda [14], patron proximity to alcohol-serving venues [19] and dim lights, dark corners, uni-sex toilets and lack of condoms in venues in South Africa [20], as well as pro-alcohol consumption social norms [21], are associated with increased HIV risk or risk behaviors. These risk behaviors include increased number of sexual partners, unprotected sex and transactional sex.[10] In addition, methodologies, such as the Priorities for Local AIDS Control Efforts (PLACE) method, have consistently found that alcohol-serving venues represent sites where people meet new sexual partners.[22] Our study adds to this literature by conducting comprehensive mapping and characterization of all alcohol-serving venues and recruitment from consenting venues (>99% of mapped venues), followed by HIV screening with assessment of care status among persons with HIV, and self-assessed HIV risk among persons without HIV, across eight communities in rural Kenya and Uganda.

Our results suggest that larger alcohol-serving venues – both in terms of patrons, workers and rooms – as well as alcohol type (commercial vs locally-produced), indoor vs outdoor spaces, and the presence of condoms and rooms for sex work, are all relevant venue attributes to consider when prioritizing outreach for HIV treatment and prevention interventions in rural Kenya and Uganda. Larger venues may also offer greater cost efficiencies, in terms of reaching an absolute number of persons with untreated HIV or increased HIV risk per venue visit, and costing analyses for the parent trial in which this study was conducted are underway. The finding that the availability of condoms on-site at venues was associated with increased HIV risk among persons without HIV, may reflect the choice by venue owners/managers to provide condoms at sites with higher risk sexual activity among patrons or workers. In contrast, we found a negative association between condom availability and the yield of persons with known HIV who were out-of-care. Whether this finding reflects low health care access as a shared characteristic of these sites and clientele – both in terms of condom availability and HIV care – due to distance from health centers, socio-economic differences or other distinct attributes of these venues, merits further exploration. Overall, less than a fifth of the venues had condoms available on site, highlighting a missed opportunity and a need for more condom availability for patrons and workers at alcohol-serving venues, in addition to referral to biomedical prevention with PrEP and PEP services.

Our study also identified venue characteristics that were associated with lower likelihood of linkage for HIV screening after recruitment and several of these characteristics were also associated with a higher yield of persons with newly diagnosed HIV or increased HIV risk, such as larger venues (greater number of rooms, patrons and workers), venues with rooms for sex work, serving commercial (as opposed to local) alcohol only, and having condoms on-site. This finding suggests that alcohol-serving venues with these characteristics may require additional recruitment strategies or HIV screening at or near the venues to fully engage a high-risk population of patrons and workers. Encouragingly, over 80% of people reached at alcohol-serving venues, including patrons and workers, linked for free clinic-based HIV screening and referral to either ART or biomedical HIV prevention. The high proportion of persons who linked for HIV screening may be due to a combination of factors, including brief HIV health messaging at the venues during recruitment, direct referral for free screening at local health facilities, the provision of multi-disease services in a subset (the intervention arm of the parent, OPAL-East Africa trial) and provision of a one-time transport reimbursement. The parent trial’s approach to alcohol-serving venue outreach offers a novel strategy for reaching people at risk of HIV or with untreated HIV outside of clinics with HIV status neutral engagement in clinic-based HIV screening and immediate referral to either biomedical HIV prevention or HIV treatment.

Our study has limitations. First, our study relied on a survey with prespecified venue characteristics, and as such may have missed certain characteristics that impact HIV screening yield. However, there is no standard survey tool for characterizing alcohol-serving venues, and the survey covered a range of venue attributes, including type of alcohol served, physical spaces (e.g., indoor vs outdoor, number of rooms, rooms for on-site sex work), availability of condoms and numbers of patrons and workers at different times during the week. Second, we relied on self-report to classify persons as at increased risk of HIV and persons with HIV as newly-diagnosed or out-of-care. Though some misclassification is possible, to our knowledge our study is among the first to evaluate associations between alcohol-serving venue characteristics and HIV screening outcomes. Lastly, our findings may be context-specific, limiting generalizability to alcohol-serving venues outside of a rural East African setting. However, the study was conducted in nearly 500 venues across eight rural communities in Kenya and Uganda and included both formal and informal drinking venues, which may improve the generalizability of our findings to similar settings and populations.

## Conclusion

In this study that mapped and characterized alcohol-serving venues in eight communities in rural Kenya and Uganda, and then recruited patrons and workers from these venues for clinic-based HIV screening, we identified venue characteristics that were associated with HIV screening outcomes. Over 80% of persons recruited from 480 venues linked to clinic for HIV screening, half of whom were either living with HIV or reported increased risk of HIV. Having a greater number of patrons was significantly associated with higher yield of persons with newly-diagnosed HIV. Having a greater number of patrons, greater number of barmaids and condoms available on site, were significantly associated with higher yield of persons at increased HIV risk. In contrast, having condoms available on site, was significantly associated with decreased yield of persons with known HIV who were out-of-care. Overall, our findings suggest that larger venues, and those with rooms for sex work and with condoms available on site, should be prioritized for enhanced, community-based interventions, both to increase HIV screening engagement and identify patrons/workers in need of HIV prevention and treatment in East Africa. These findings may aid programs in targeting certain types of alcohol-serving venues during community-based outreach for HIV testing, prevention and treatment services.

## Data Availability

A complete de-identified participant-level dataset sufficient to reproduce the study findings will be made available approximately 1 year after completion of the ongoing trial ( NCT05862857), following approval of a concept sheet summarising the analysis to be performed. Further inquires can be directed to the Principal Investigator at

## Acknowledgments

We gratefully acknowledge the Kenya Ministry of Health, Uganda Ministry of Health, the research and administrative teams in San Francisco, Kenya, and Uganda, our collaborators and all of the communities and participants involved in the study.

## Supporting information

**S1 Table. Venue characteristics associated with likelihood of linking to facility-based screening among participants who received recruitment cards; unadjusted odds ratios.**

**S2 Table. Univariate (unadjusted) associations between alcohol-serving venue characteristics and HIV screening outcomes among adults recruited from alcohol-serving venues, including: a) newly diagnosed HIV, b) known HIV and out-of-care, and c) increased risk of HIV, if HIV uninfected.**

